# Efficacy and Tolerability of Lesion Network Guided Transcranial Electrical Stimulation in Outpatients with Psychosis Spectrum Illness: A Nonrandomized Controlled Trial

**DOI:** 10.1101/2023.03.31.23287980

**Authors:** Nicolas Raymond, Robert M.G. Reinhart, Rebekah Trotti, David Parker, Shrey Grover, Bilge Turkozer, Dean Sabatinelli, Rachal Hegde, Deepthi Bannai, Swetha Gandu, Brett Clementz, Matcheri Keshavan, Paulo Lizano

**Affiliations:** Department of Psychiatry, Beth Israel Deaconess Medical Center, Boston, MA, USA; Department of Psychological and Brain Science, Boston University, Boston, MA, USA; Department of Human Genetics, Emory University School of Medicine, Atlanta, GA, USA; Department of Psychiatry, Division of Child and Adolescent Psychiatry, Massachusetts General Hospital and McLean Hospital, MA, USA; Department of Psychology, University of Georgia, Athens, GA, USA; Department of Psychiatry, Harvard Medical School, Boston, MA, USA; Division of Translational Neuroscience, Beth Israel Deaconess Medical Center, Boston, MA, USA

**Keywords:** Positive Symptoms, Transcranial Electrical Stimulation, Lesion Network Mapping

## Abstract

**Importance:** Transcranial electrical stimulation (tES) may improve psychosis symptoms, but few investigations have targeted brain regions causally linked to psychosis symptoms. We implemented a novel montage targeting the extrastriate visual cortex (eVC) previously identified by lesion network mapping in the manifestation of visual hallucinations.

**Objective:** To determine if lesion network guided HD-tES to the eVC is safe and efficacious in reducing symptoms related to psychosis.

**Design, Setting, and Participants:** Single-center, nonrandomized, single-blind trial using a crossover design conducted in two 4-week phases beginning November 2020, and ending January 2022. Participants were adults 18-55 years of age with a diagnosis of schizophrenia, schizoaffective or psychotic bipolar disorder as confirmed by the Structured Clinical Interview for DSM-V, without an antipsychotic medication change for at least 4 weeks. A total of 8 participants consented and 6 participants enrolled. Significance threshold set to <0.1 due to small sample size.

**Interventions:** 6 Participants first received HD-tDCS (direct current), followed by 4 weeks of wash out, then 4 received 2Hz HD-tACS (alternating current). Participants received 5 consecutive days of daily (2 × 20min) stimulation applied bilaterally to the eVC.

**Main Outcomes and Measures:** Primary outcomes included the Positive and Negative Syndrome Scale (PANSS) total, positive, negative, and general scores, biological motion task, and Event Related Potential (ERP) measures obtained from a steady state visual evoked potential (SSVEP) task across each 4-week phase. Secondary outcomes included the Montgomery-Asperg Depression Rating Scale (MADRS), Global Assessment of Functioning (GAF), velocity discrimination task, visual working memory task, and emotional ERP across each 4-week phase.

**Results:** HD-tDCS improved general psychopathology in the short-term (d=0.47; p_fdr_=0.03), with long-term improvements in general psychopathology (d=0.62; p_fdr_=0.05) and GAF (d=-0.56; p_fdr_=0.04) with HD-tACS. HD-tDCS reduced SSVEP P1 (d=0.25; p_fdr_=0.005), which correlated with general psychopathology (β=0.274, t=3.59, p=0.04). No significant differences in safety or tolerability measures were identified.

**Conclusions and Relevance:** Lesion network guided HD-tES to the eVC is a safe, efficacious, and promising approach for reducing general psychopathology via changes in neuroplasticity. These results highlight the need for larger clinical trials implementing novel targeting methodologies for the treatments of psychosis.

**Trial Registration:** ClinicalTrials.gov Identifier: NCT04870710

**Key Points:** *Question:* Is lesion network guided neurostimulation an efficacious, safe, and targeted approach for treating psychosis?

*Findings:* In this single-center, nonrandomized, crossover, single-blind trial of 6 outpatients with psychosis, improvement in general psychopathology was seen in the short-term with HD-tDCS (high-definition transcranial direct current stimulation) and long-term with HD-tACS (alternating current) targeting the extrastriate visual cortex (eVC). HD-tDCS reduced early visual evoked responses which linked to general psychopathology improvements. Overall, both stimulations were well tolerated.

*Meaning:* Study findings suggest that lesion network guided HD-tES to the eVC is a safe, efficacious, and promising approach for reducing general psychopathology via neuroplastic changes.

## Introduction

Transcranial electrical stimulation (tES) modulates cortical activity and influences cognition^1^, perception^2^, and positive symptoms in psychosis^3^. Few researchers have integrated recent neuroimaging findings to identify optimal stimulation targets, such as location, frequency, and circuits^4^. Innovations in tES hardware and software now allows for more focal stimulation (using high definition tES, HD-tES) compared to sponge montages^5^ and greater spatial target engagement using current flow models^6^. While HD-tES advances have been effective for the treatment of neuropsychiatric disorders^7^ few studies have used HD-tES in psychosis^1,4,8,9^.

Psychotic disorders consist of negative symptoms^10^, positive symptoms^11^ and cognitive deficits^12^. Positive symptoms, such as hallucinations are often debilitating with visual hallucinations (VH) associated with more severe morbidity, delusions, suicidal behavior, and catatonia^13^. While antipsychotics treat positive symptoms, ~30% of individuals are treatment resistant^14^, which may result in metabolic dysregulation^15^, agranulocytosis, and risk of seizures^16^. Thus, there is a critical need for novel, neurobiologically informed, non-invasive, and safe treatments for psychosis symptom management, such as HD-tES.

To optimize tES parameters we used a combination of neuroimaging, neurophysiological, and cause-effect studies. The extrastriate visual cortex (eVC) was of particular importance due to its role in motion perception, neurocognition, and social cognition^17,18^. For instance, in a large cross-sectional neuroimaging study we identified thinning of the eVC (V5/MT) across the psychosis spectrum compared to controls, which correlated with poor cognition and response inhibition^19^. In fMRI studies examining active visual and/or auditory hallucinations in drug-free adolescents with brief psychotic disorders or adults with psychosis spectrum disorders, the authors found activation of the primary and secondary visual cortices^20,21^. Results from a lesion networking mapping (LNM) study, a powerful tool used to make causal inferences from lesions causally linked to symptoms^22^, identified the eVC to be implicated in VH^23^. Pathologically elevated eVC activity has also been demonstrated in psychotic^24^. Lastly, a study examining the neural basis of motion perception in schizophrenia found that reduced V5/MT activation was associated with lower delta (2Hz) evoked amplitude during motion related tasks and poorer cognitive performance^25^. This convergent body of work highlights the importance of the eVC and delta frequency in psychosis and provides a framework for neurobiologically informed treatment with HD-tES.

To examine the translational value of the eVC in psychosis, we conducted a proof-of-concept single blind crossover study at a single site to characterize the efficacy and safety of using cathodal HD-tDCS (transcranial direct current stimulation) or delta frequency (2hz) HD-tACS (transcranial alternating current stimulation) in improving psychosis symptoms, visual processing, and visual evoked potentials.

## Methods

### Participants

This study enrolled outpatients beginning October 1, 2020 with the final study visit completed on January 2, 2022. This study was approved by the Institutional Review Board at Beth Israel Deaconess Medical Center, Massachusetts. Participants signed written informed consent and were compensated for their participation (**see trial protocol in Supplement 1**).

We intended to recruit 10 individuals (5 sham and 5 HD-tDCS) between the ages of 18 to 55 years with schizophrenia, schizoaffective disorder, or psychotic bipolar disorder using the Structured Clinical Interview for DSM-V, and with a lifetime history of VH and/or experiencing mild to moderate symptoms of VH. Since recruitment efforts were hindered due to institutional restrictions during the COVID-19 pandemic, we removed the VH requirement and sham condition. Instead, the study was transitioned to a crossover design using HD-tDCS followed by 2Hz HD-tACS.

Participants had no antipsychotic medication change in the month prior to participation. Participants were excluded if they had an intelligence quotient <60, any major medical or neurologic condition, a diagnosis of substance abuse or positive urine drug screen, history of moderate-to-severe visual impairment secondary to glaucoma, cataract or macular degeneration, serious medical illness or instability requiring hospitalization within the last year, relevant skin allergies, metallic or electronic implants, or if they were pregnant or breastfeeding.

### Procedure

This proof-of-concept study used a between-participants, single blind, non-randomized, crossover design, with two tES treatment conditions. Participants first received HD-tDCS, followed by 4 weeks of wash out, then received 2Hz HD-tACS (**Figure 1A)**. Clinical assessments were performed by a psychiatrist at baseline, day 5 and 1-month. Participants arrived at the hospital on a Monday, were briefed on study procedures by a research assistant, followed by electroencephalography (EEG) including a steady-state visual evoked potential (SSVEP) task, and emotional scene processing task (International Affective Picture System; IAPS). Visual processing tasks were conducted while seated in a dark room under the supervision of study staff (**Figure 1B)**. Then, 2 sessions of 20 min HD-tDCS was administered daily for 5 days while the participant sat comfortably, quietly and without disruption. A 15-20 min break was provided between the 2 sessions and participants were asked to complete a brief sensation questionnaire related to sensations felt during the administration of tES. On a Friday, and after 5 days of treatment, baseline assessments were repeated. These assessments were performed again after 1-month. Participants then received HD-tACS, which consisted of the same study procedures as HD-tDCS.

**Figure 1:**
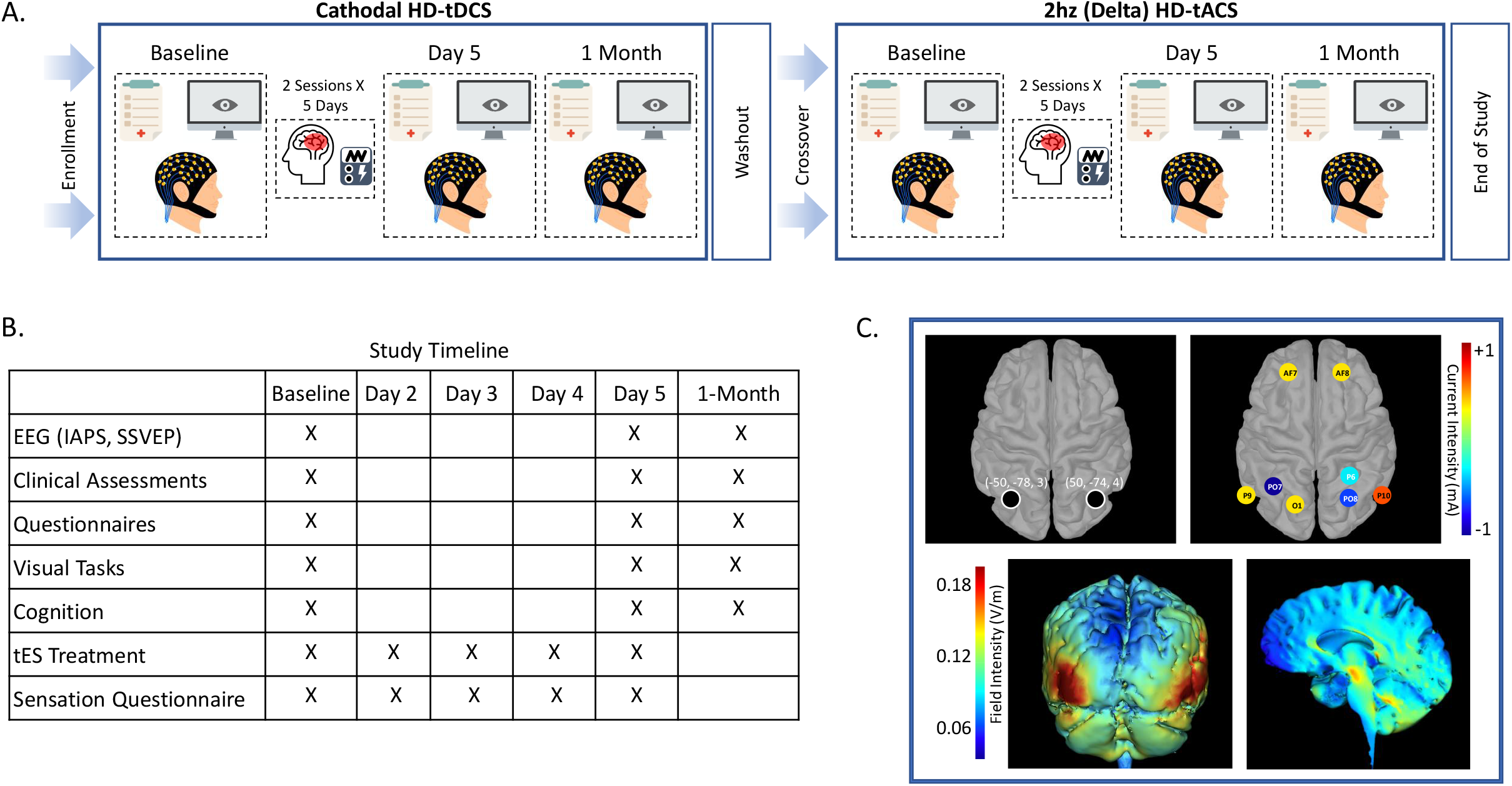
Study Design, Timeline and Transcranial Electrical Stimulation (tES) Montage: A. Depicts the experimental crossover study design. B. Demonstrates the study timeline showing when the primary and secondary outcomes were collected, as well as the days participants received electrical stimulation. C. Shows the stimulation coordinates in Montreal Neurologic Institute (MNI) space for the bilateral extrastriate visual cortex target, stimulation electrode montage (current intensity depicted in heatmap), and the current flow modeling (field intensity depicted in heatmap). **Note**: HD-tDCS, High-Definition Transcranial Direct Current Stimulation; HD-tACS, HHD-Transcranial Alternating Current Stimulation; EEG, Electroencephalogram; IAPS, International Affective Picture System; SSVEP, Steady State Visual Evoked Potential;

### Treatment

HD-tDCS and HD-tACS was delivered by a Soterix MXN-9 High Definition-Transcranial Electrical Current Stimulator, Model 9002A (**Supplement 2**). The stimulation montage was designed to target the lesion network mapping findings associated with VH, which identified the bilateral eVC^23^ (**Figure 1C)**. The delta (2Hz) frequency peak for this study was extracted from the Maritnez et al 2018 paper, which conducted a time-frequency analysis of a motion processing task in patients with schizophrenia (**Supplement 3**). Electrical current field modeling^6^ using HD-Explore and HD-Targets (Soterix Medical) guided decision-making about where to place electrodes, with the goal of delivering focalized current to the bilateral eVC. The montage consisted of cathodal PO7 and anodal P9, O1, AF7 on the left, and cathodal P6, P08 and anodal P10, AF8 on the right according to the International 10-10 System. HD-tACS used the same montage but with 2hz in-phase alternating current being delivered (**Figure 1C)**.

### Outcome Measures

The primary outcomes examined were the Positive and Negative Syndrome Scale (PANSS), biological motion detection, and SSVEP between timepoints and stimulation montages. PANSS total, positive, negative, and general scores were used. Visual processing outcomes were obtained by a biological motion task to assess the accuracy for determining the direction of motion^26^ (**Supplement 4**). Event Related Potential (ERP) measures were obtained through a SSVEP task to assess changes in biomarkers of the early visual response, the P1 and N1 (**Supplement 5**).

The secondary outcomes examined included the Montgomery-Asberg Depression Rating Scale (MADRS), Global Assessment of Functioning (GAF), visual processing behavioral tasks, and emotional processing ERPs. Visual processing measures were obtained through a velocity discrimination and a visuospatial working memory task to assess accuracy of speed detection and visual working memory, respectively^26^ (**Supplementary 4**). Emotional ERP measures were obtained using the IAPS, which consists of unpleasant, pleasant, and neutral scene stimuli, to assess changes in a motivationally-relevant early visual biomarker, the early posterior negativity (EPN)^27^ (**Supplementary 5**).

Exploratory analyses included determining whether significant (p<0.1) target engagement of EEG measures using tES would be correlated with significant (p<0.1) changes in clinical or behavioral measures.

### Statistical Analysis

All statistics were performed using R software (v4.1.2) and RStudio. For individuals missing 1-month assessments (HD-tDCS n=1, HD-tACS n=1), values were imputed using the Amelia package^28^ while accounting for scores across sessions. The “ggstatsplot” package was used for statistical analysis and plots^29^. The “WRS2” package was used for two-way ANOVA^30^.

Chlorpromazine equivalents was calculated using “chlorpromazineR” and the Leucht et al methodology^31^. We used non-parametric tests consisting of the Friedman and Durbin Conover tests to examine within group differences. Trimmed means two-way ANOVA models were used to examine group (HD-tDCS, HD-tACS) by session (baseline, day 5 & 1-month) interactions. To assess the relationship between changes (follow up - baseline) in clinical and EEG measurements, rank-based estimation regression while controlling for skewness^32^ was used with baseline clinical measurements used as a covariate. An alpha value of 0.10 was set for significance due to the sample size of the study and to help identify effect sizes to power future large scale trials^33^. Kendall (*W*) and Rank Biserial Effect Size (RBES) was calculated. Corrected p-values are reported for pairwise comparisons. To confirm significant results analyses were re-run using the non-imputed dataset.

## Results

A total of 6 participants with a psychosis spectrum disorder were enrolled in the study. All 6 received HD-tDCS and 4 received 2Hz HD-tACS (**Figure 2**). Baseline demographic and clinical characteristics are summarized in **Table 1**.

**Figure 2.**
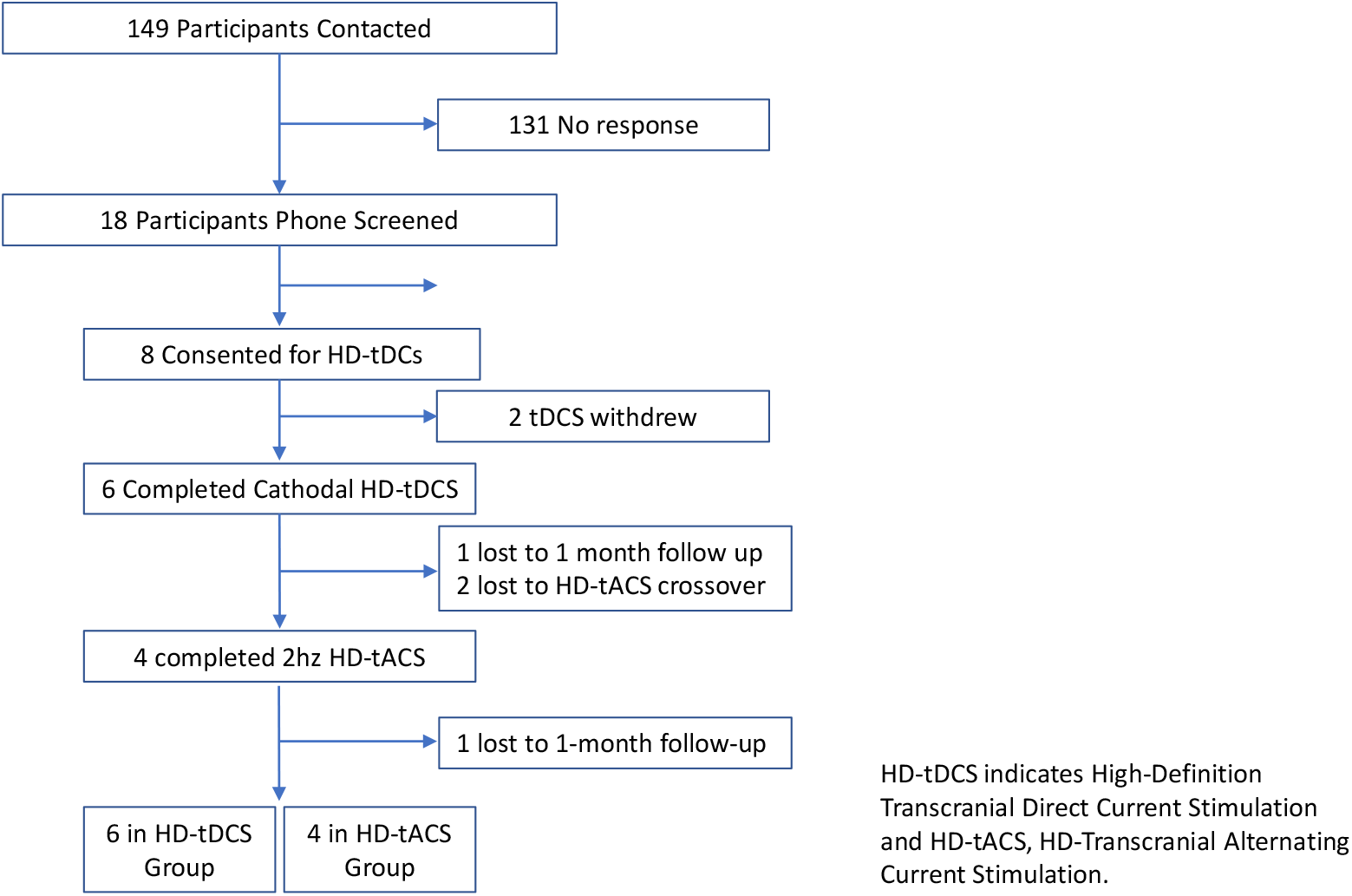
CONSORT Flow Diagram

**Table 1:**
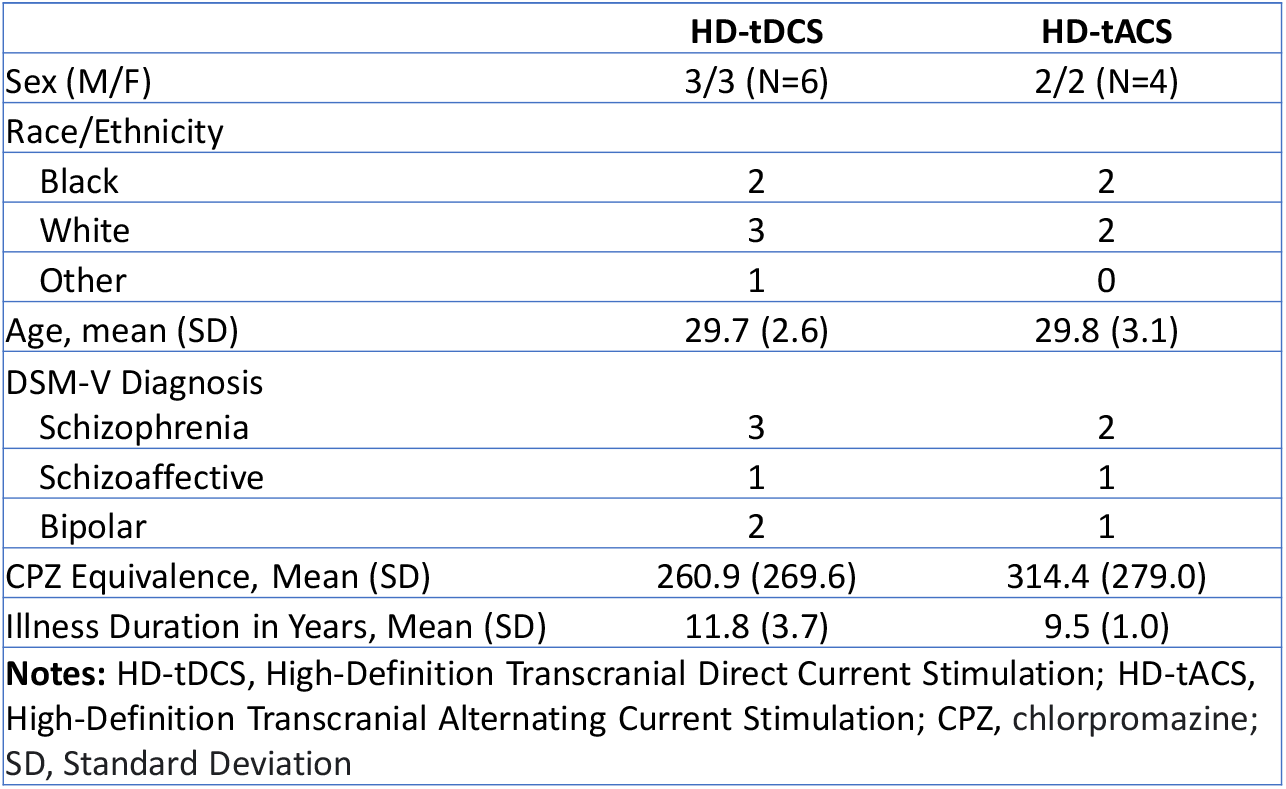
Baseline Demographic Characteristics

|  | **HD-tDCS** | **HD-tACS** |
| --- | --- | --- |
| Sex (M/F) | 3/3 (N=6) | 2/2 (N=4) |
| Race/Ethnicity |  |  |
| Black | 2 | 2 |
| White | 3 | 2 |
| Other | 1 | 0 |
| Age, mean (SD) | 29.7 (2.6) | 29.8 (3.1) |
| DSM-V Diagnosis |  |  |
| Schizophrenia | 3 | 2 |
| Schizoaffective | 1 | 1 |
| Bipolar | 2 | 1 |
| CPZ Equivalence, Mean (SD) | 260.9 (269.6) | 314.4 (279.0) |
| Illness Duration in Years, Mean (SD) | 11.8 (3.7) | 9.5 (1.0) |
| **Notes:** HD-tDCS, High-Definition Transcranial Direct Current Stimulation; HD-tACS, High-Definition Transcranial Alternating Current Stimulation; CPZ, chlorpromazine; SD, Standard Deviation |  |  |

### Primary Outcomes

There were significant differences across sessions for PANSS general symptoms in the HD-tDCS (*W*=0.42; p=0.04) and HD-tACS condition (*W*=0.58; p=0.07), but not for total, positive or negative symptoms (**Table 2A, Figure 3A**). Post hoc comparisons in the HD-tDCS showed a significant reduction from baseline to day 5 for PANSS general scores (RBES=0.47; p_fdr_=0.03) and significant increase from day 5 to 1-month (RBES=-0.50; p_fdr_=0.03). For HD-tACS, significant reductions in PANSS general score between day 5 and 1-month (RBES=0.69; p_fdr_=0.05) and from baseline to 1-month (RBES=0.62; p_fdr_=0.05) was observed. There were no significant differneces between HD-tDCS 1-month and HD-tACS baseline nor between HD-tDCS baseline and HD-tACS 1-month (**eFigure 1**). These analyses were repeated without imputed data and results were similar for the HD-tDCS and HD-tACS findings (**Supplement 6**). Post hoc analysis showed a significant group by session interaction (F=12.42, p=0.02) between HD-tDCS and HD-tACS (**eTable 1, Figure 3B**).

**Table 2A.**
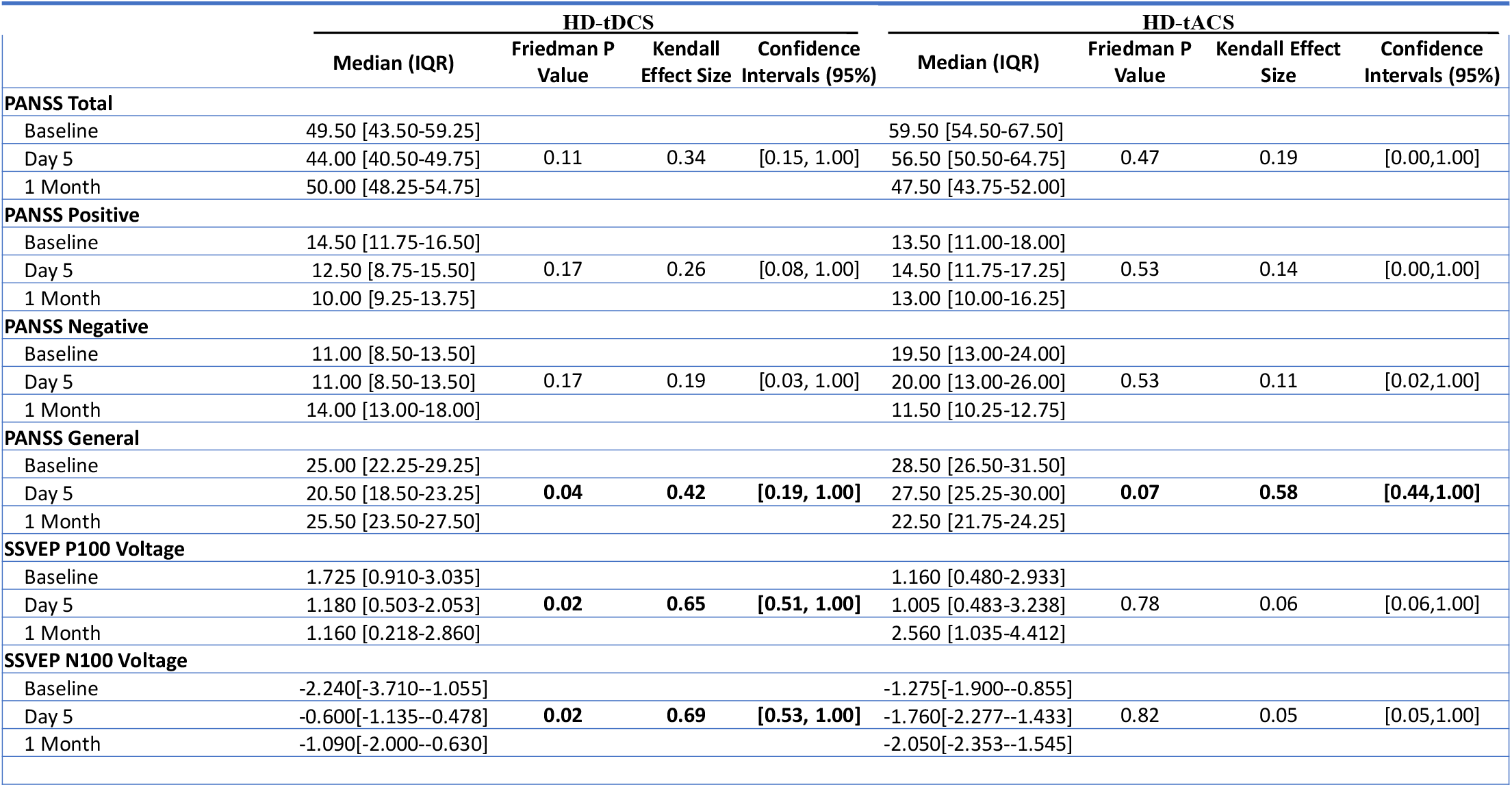
Primary Outcome Results

**Figure 3:**
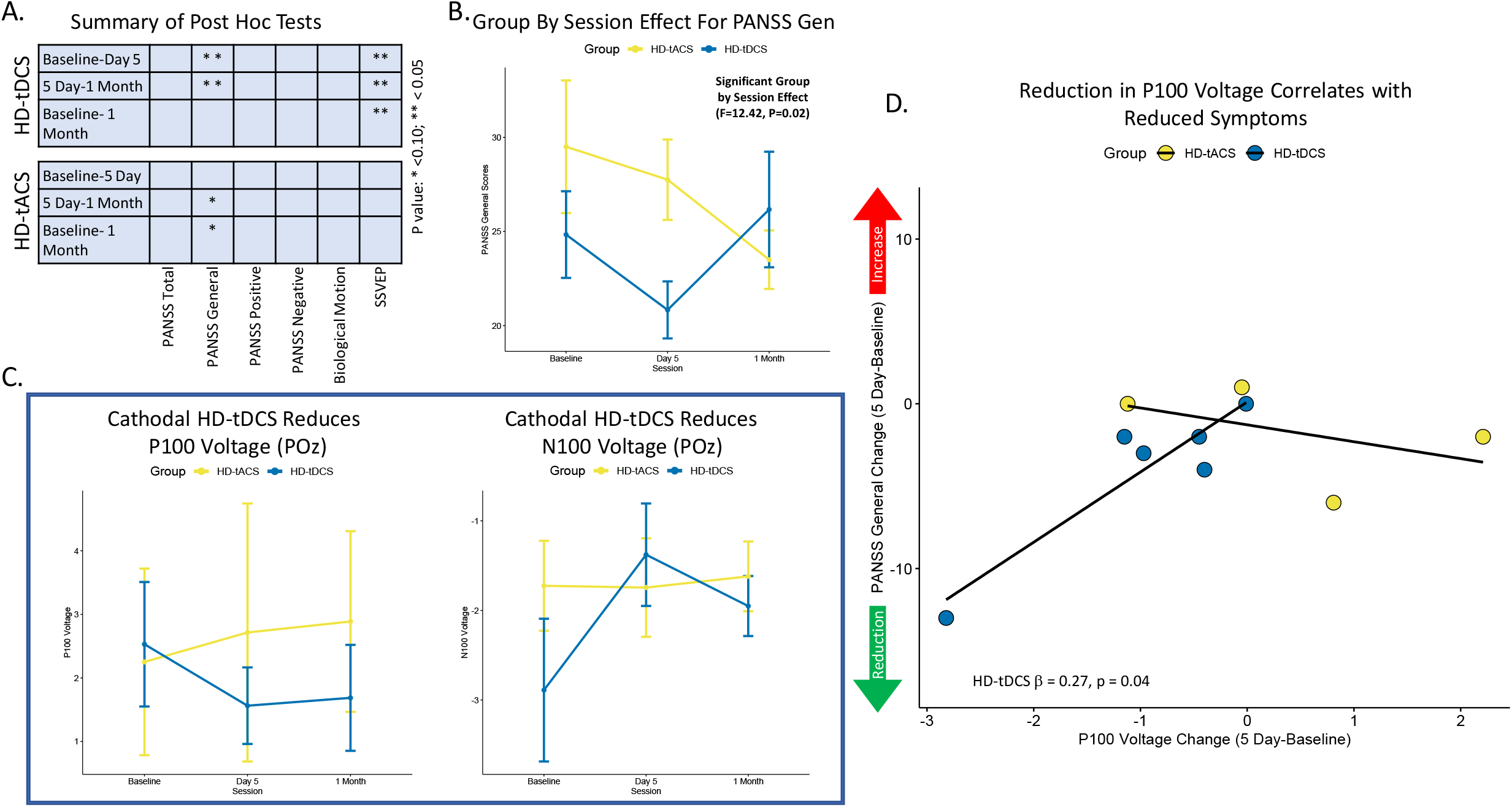
Primary Outcome Results: A. Demonstrates the summary of post-hoc pairwise comparisons by session contrasts for both HD-tDCS and HD-tACS. B. Depicts the group by session interaction effect for the PANSS General score. C. Shows the SSVEP P100 and N100 results at the Poz sensor across sessions. D. Demonstrates the regression results between change scores (5 Day-Baseline) for P100 Voltage and PANSS General score with a significant result in the HD-tDCS condition. **Notes:** High-Definition Transcranial Current Stimulation; HD-tACS, High-Definition Transcranial Alternating Current Stimulation; PANSS, Positive and Negative Syndrome Scale; SSVEP, Steady State Visual Evoked Potential

There were significant differences across sessions for the SSVEP P1 voltage in the HD-tDCS group for bilateral trials at POz (*W*=0.65; p=0.02) (**Table 2A, Figure 3A,C**). HD-tDCS post hoc analyses showed a significant decrease in voltage for P1 from baseline to 5 day (RBES=0.25; p_fdr_=0.005) and baseline to 1-month (RBES=0.33; p_fdr_=0.008). The SSVEP N1 voltage was significantly different across sessions in the HD-tDCS group for bilateral POz (*W*=0.69; p=0.02). HD-tDCS post hoc analyses showed a significant increase in voltage for N1 from baseline to 5 day (RBES=-0.56; p_fdr_=0.002) and baseline to 1-month (RBES=-0.28; p_fdr_=0.04), as well as a significant decrease from 5 day to 1-month (RBES=0.39; p_fdr_=0.04). There were no significant session differences noted for P1 and N1 in the HD-tACS group. There was no significant group by session effect noted for P1 or N1 (**eTable 1, Figure 3C**). These results were repeated without imputed values and the results were similar (**Supplement 6**).

There were no significant differences observed on the biological motion task for either treatment condition (**eTable 2**).

In exploratory analyses, a significant relationship was identified between the improvement in PANSS general score and the reduction in P1 observed between day 5 and baseline (β=0.274, t=3.59, p=0.04) (**eTable 3, Figure 3D**).

### Secondary Outcomes

There were significant differences across sessions for GAF scores in the HD-tACS condition (*W*=0.44; p=0.06) (**Table 2B, Figure 4A**). Post hoc comparisons in the HD-tACS showed a significant increase in GAF from day 5 to 1-month (RBES=-0.56; p_fdr_=0.05) and baseline to 1-month (RBES=-0.56; p_fdr_=0.04). These analyses were repeated without imputed data and results were similar for the HD-tACS findings (**Supplement 6**). There was no group by session effect observed for GAF (**eTable 1, Figure 4B**). There were no significant differences noted for MADRS within or between conditions (**Table 2B, eTable 1**).

**Table 2B.**
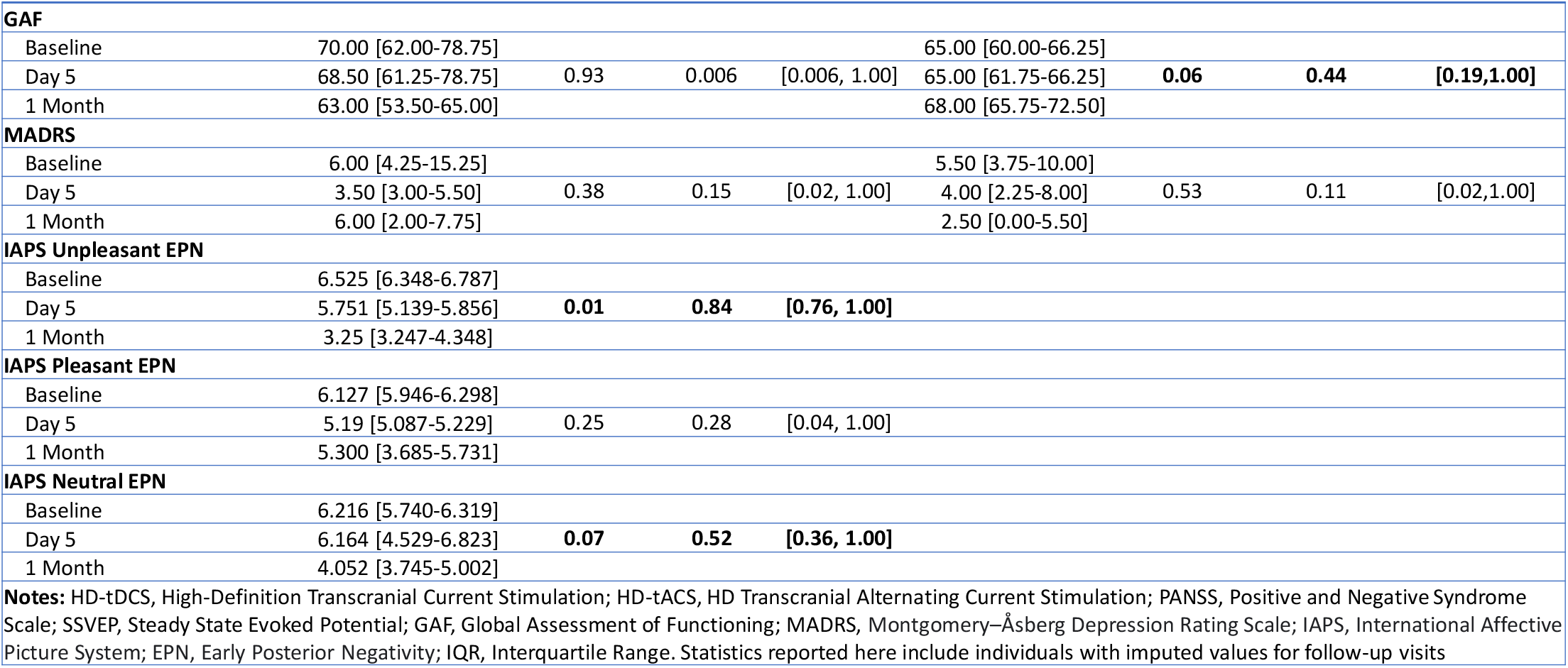
Secondary Outcome Results

**Figure 4:**
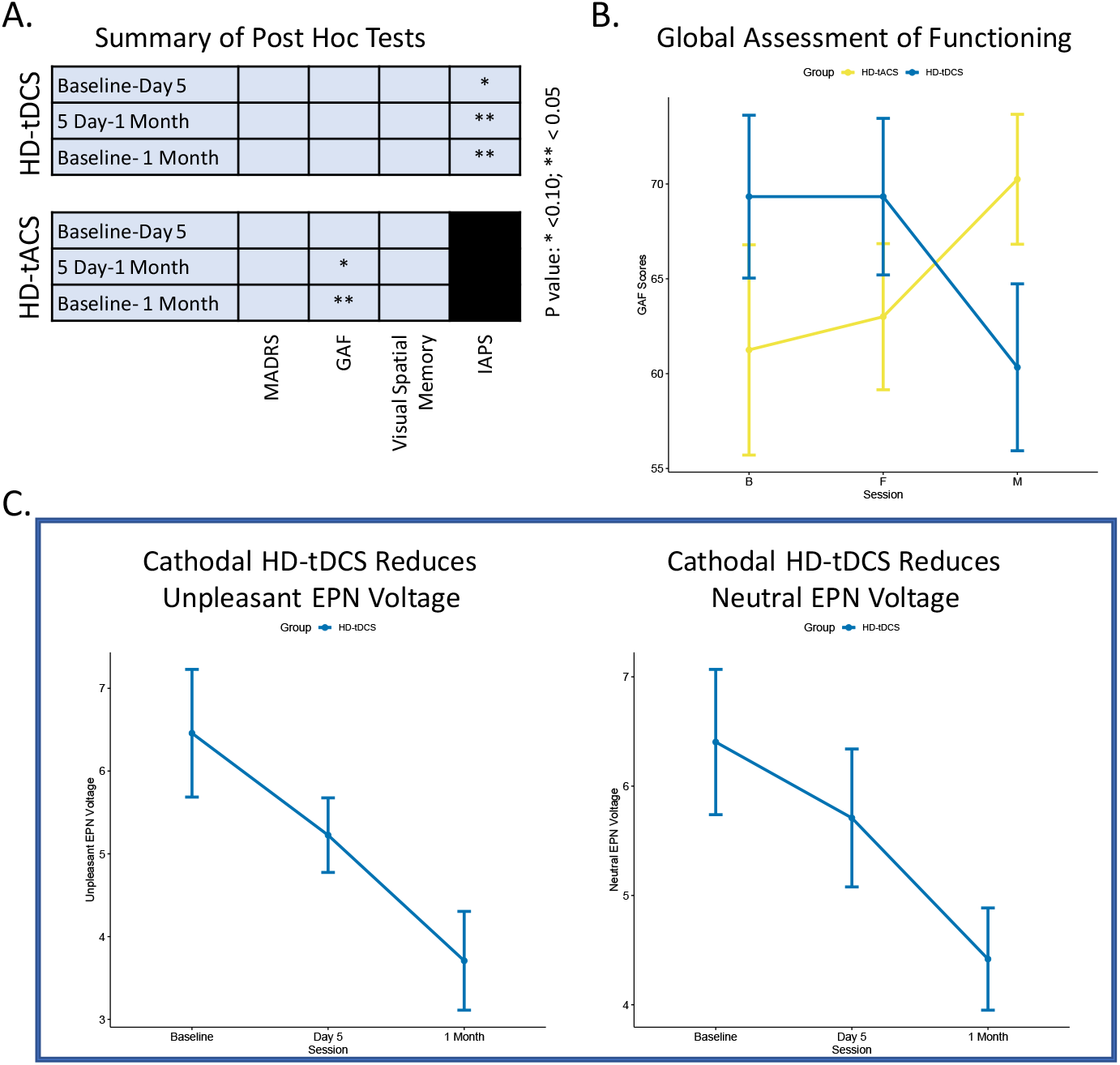
Secondary Outcome Results: A. Demonstrates the summary of post-hoc pairwise comparisons by session contrasts for both HD-tDCS and HD-tACS. B. Depicts the results for GAF scores across sessions for both HD-tDCS and HD-tACS with a significant reduction in the HD-tACS group at 1 Month. C. Shows the IAPS EPN Voltage for Unpleasant and Neutral stimuli at P6, P7, PO6, PO7, O1, and O2 sensors across sessions. **Notes:** HD-tDCS, High-Definition Transcranial Current Stimulation; HD-tACS, HD Transcranial Alternating Current Stimulation; GAF, Global Assessment of Functioning; IAPS, International Affective Picture System; EPN, Early Posterior Negativity. 1 participant in the HD-tDCS condition was not able to complete IAPS assessments.

There were significant differences across sessions for the IAPS EPN voltages in the HD-tDCS condition for both unpleasant (*W*=0.84; p=0.01) and neutral (*W*=0.52; p=0.07) stimuli, but not for pleasant (**Table 2B, Figure 4C**). Pairwise comparisons in the HD-tDCS condition showed a significant decrease in response amplitude to unpleasant stimuli from baseline to day 5 (RBES=-0.68; p_fdr_=0.07), day 5 to 1-month (RBES=0.76; p_fdr_=0.004) and baseline to 1-month (RBES=0.84; p_fdr_=0.0007). Pairwise comparisons showed that response amplitudes to neutral stimuli decreased from baseline to 1-month (RBES=0.76; p_fdr_=0.06). These analyses were repeated without imputed IAPS data and results were similar for the HD-tDCS findings in the unpleasant stimuli, but not significant for neutral stimuli (**Supplement 6**).

There were no significant differences observed on the visual spatial working memory or velocity discrimination task for either treatment condition (**eTable 2**).

In exploratory analyses, no significant relationship was identified between the improvement in PANSS general score and the reduction in unpleasant (β=0.529, t=2.18, p=0.16) or neutral (β=0.173, t=0.37, p=0.75) stimuli observed between day 5 and baseline (**eTable 3**).

There were no serious adverse events reported in either stimulation condition and no participant withdrew from the study due to side effects. The stimulation montage was well tolerated and no participant reported above a moderate sensation on the sensation scale (**eFigure 2**).

## Discussion

This is the first tES intervention for psychosis to precisely target the eVC, guided by lesion network mapping and HD-tES current flow models. We demonstrated that stimulating this region using HD-tDCS may improve general psychopathology in the short-term (5 days), with longer-term (1-month) improvements in general psychopathology and functioning noted with HD-tACS. Furthermore, eVC stimulation with HD-tDCS may induce a sustained reduction in early visual ERPs from visual steady-state and emotional scene paradigms, but this effect was not observed using HD-tACS. Regression analysis in the HD-tDCS condition indicates that general psychopathology and electrophysiological reductions are linked, suggesting that engaging the eVC with HD-tES may play a role in the alleviation of psychosis symptoms. Lastly, both HD-tES montages used in this study were well tolerated (**eFigure 2**).

The HD-tDCS general psychopathology results are consistent with findings in the literature from randomized control trials with 8 studies demonstrating short-term improvements (SMD=0.31), while 4 studies did not show longer-term benefits at 4-12 weeks (SMD=0.15)^34^. These studies used 2mA stimulation intensity, anodal to the left dorsolateral prefrontal cortex (F3) and cathodal to right frontal (F4) or left temporoparietal junction (T3, P3), stimulation area ranged from 25-35cm^2^, and sessions ranged from 5-10 sessions. Further support comes from a case report of a patient with treatment resistant auditory hallucinations and VH who underwent cathodal tDCS to Oz for 10 sessions and then the temporoparietal area for 10 sessions, and they experienced a 29% reduction in general psychopathology symptoms at 1-month^35^. The HD-tACS general psychopathology findings are also consistent with a case series of 3 clozapine resistant patients with schizophrenia receiving theta (4.5 Hz) tACS demonstrating an 18% improvement in symptoms^36^. This study used 2 mA stimulation intensity, F3 and F4 electrode placement, 25cm^2^ area, for 20 sessions over 4 weeks. While these studies are promising they were conducted using sponge montages, which decrease the focality of stimulation, and traditional montages were used targeting primarily frontal, temporal, and parietal regions, which don’t specifically target networks associated with behavior or psychosis symptomatology. Our study expands on this literature by demonstrating that HD-tDCS to the eVC which is causally linked to VH^23^ and motion processing^25^, resulted in a larger short-term effects size change (RBES=0.47) for general psychopathology than has been reported previously. We are also the first to demonstrate that 2Hz tACS to the eVC can result in a long-term moderate effect size (RBES=0.62) improvement at 1-month, which may be due to neuroplastic changes induced by phase locking of intrinsic brain rhythms^37^, but further work is needed in this area.

The mechanism through which HD-tDCS or HD-tACS decreases general psychopathology is not fully understood. However, the findings of the present study suggest that HD-tDCS to the eVC induces a neuroplastic change to the SSVEP P1 and IAPS EPN ERPs with the former being correlated with a change in general psychopathology, however, this effect was not observed with HD-tACS. This observation may be explained by the fact that tDCS can modulate cortical excitability using anodal stimulation which tends to increase (i.e. the resting potential becomes less negative), while cathodal stimulation tends to decrease the underlying membrane potential (i.e. the resting potential becomes more negative) ^38,39^. Furthermore, studies have demonstrated that tDCS can modulate visual cortical function in a polarity-dependent manner, where anodal stimulation can increase and cathodal stimulation can decrease the amplitude of the N70 component from the visual-evoked potential ^40^. While there is no study to date examining the relationship between P1 and general psychopathology, a study using dynamic facial expressions to examine ERP responses in schizophrenia, found that greater N200 latency was associated with lower general psychopathology scores^41^. Different from tDCS, tACS is known to modulate endogenous neural oscillations by applying oscillating electrical current with a periodic waveform to the brain^42^. Using tACS to target the occipital cortex, it was demonstrated that different stimulation frequencies can interact with endogenous rhythmic activities in a frequency-specific manner to induce phosphenes^43^. While these studies are informative, more research is needed to better understand the mechanisms underlying the improvement in general psychopathology.

## Limitations

We acknowledge several important limitations in understanding our results. First, the sample size was small despite the strength of a cross-over design. Second, imputed data was used for 1-month assessments, but the results were similar when repeated using unimputed data. Third, subjects were stable outpatients not experiencing clinically significant symptoms and future studies should be performed in an acute population. Fourth, velocity discrimination measurements could not be used in our study due to poor data quality, and this metric is likely a better behavioral target than biological motion when stimulating the eVC^44^. Additionally, the lack of change in biological motion scores from the two stimulations arms suggest that this task may be a reliable way to measure the absence of off target effects. Fifth, the lack of positive psychosis symptom findings may be due to a lack of self-reported psychosis symptoms scales, which may be a more accurate measure of predicting outcomes^45,46^. Lastly, we did not use each individuals structural MRI, which would have allowed us to personalize the stimulation location and current flow^47,48^, as well as maximize the effects of HD-tES. Despite these limitations, this is an important proof of concept study that lays the foundation for future studies investigating the treatment of positive and general symptoms of psychosis with HD-tES.

## Conclusions

Findings from the present study suggest that lesion network guided HD-tES to the eVC is a safe, efficacious, and promising approach for reducing general psychopathology via changes in neuroplasticity. These results highlight the need for larger clinical trials implementing novel targeting methodologies and montages with the hopes of identifying effective future treatments for psychosis.

## Data Availability

All data produced in the present study are available upon reasonable request to the authors

## SUPPLEMENTARY FILES

### Supplement 1

See attached trial protocol

### Supplement 2

#### Stimulation Setup and Montage Modeling

Both stimulation procedures (HD-tDCS & HD-tACS) used 12-mm-diameter Ag-Agl electrodes placed in high-definition electrode holders provide through Soterix Medical. Additionally, electrode holders were filled with high conductive gel. All electrical field modeling was performed on HD-Explore and HD-Targets (Soterix Medical). Modeling was preformed to help guide electrode placement and adjustments were made to deliver maximum focalized current to MNI coordinates (−50, -78, 30) & (50, -74, 4). All modeling was performed on a standardized brain included in Soterix Medical software. Overall current intensity (peak to baseline) was set to 2mA.

#### HD-tDCS

The direct current stimulation parameters including sensor location and current intensity are as described: AF7, 0.333 mA; AF8, 0.35 mA; P6, -0.325 mA; P9, 0.333 mA; P10, 0.65 mA; PO7, - 1.00 mA; PO8, -0.675; O1, 0.334 mA.

#### HD-tACS

Stimulation parameters including sensor location and current intensity are as described: AF7, 0.333 mA; AF8, 0.35 mA; P6, -0.325 mA; P9, 0.333 mA; P10, 0.65 mA; PO7, -1.00 mA; PO8, - 0.675; O1, 0.334 mA. A bipolar sinusoidal in-phase alternating current was delivered at 2Hz.

#### Stimulation Procedure

All stimulation procedures included a 30 second ramp up and ramp down. Stimulation lasted 20min and was delivered twice daily for five days. A short break (15min) took place in-between daily stimulations. During the break, participants were asked to fill out the sensation questionnaire. After the second stimulation was completed, participants again filled out the sensation questionnaire. All participants were seated comfortably and were asked to remain awake but not to engage in any tasks or behaviors during the stimulation.

### Supplement 3

We extracted the peak delta frequency from the grand average power plot in the Martinez et al 2018 paper. We used WebPlotDigitizer to estimate the delta power peaks and found the peak of 2.15 Hz for the adult healthy control group and 1.8 Hz for the young healthy control group.

These values reflect the center of the darkest area in the peaks. There was also a very weak peak visible in the SZ group that came to about 2.43 Hz. The attenuated psychosis group did not have a discernable delta activity peak that could be easily measured. Based on these estimates, we concluded that it was reasonable to entrain the SZ population at the average frequency of the healthy groups, namely 2 Hz.

### Supplement 4

#### Evaluation of Visual Perceptual Functions using Behavioral Tasks

All experimental stimuli presented on an CRT monitor. Participants’ head stabilized using a chin and head rest at a distance of 75 cm from the screen. Participants were asked to view the stimuli and indicate the answer of the directed question using a keyboard.

#### Biological Motion

Biological motion perception was assessed using point-light animations (12 dots on the head and major joints of the body) walking either rightward or leftward^49^. The target animation was embedded in a number of random-moving noise dots (24, 48, or 72) to manipulate the difficulty level of the task. Participants were asked to indicate the direction that the animation was walking towards. The task lasted for about 4 minutes.

#### Visuospatial Working Memory

Visuospatial working memory was evaluated using the “Odd one out” task ^50^. Participants were asked to identify and remember the location of the odd shape out of three shapes. The procedure was repeated with three new shapes following an interval. Participants were asked to respond by indicating where the odd shapes appeared, in the correct order of appearance. Two correct trials on each level led to progression to the next level where the item load was increased by one. The session was terminated when two trials on the same level were incorrect. The final score was calculated based on the performance on the highest level achieved where at least one trial was passed. This task lasted for 3-5 minutes.

#### Velocity Discrimination

Velocity discrimination performance was determined using constant stimuli method. Participants asked to indicate the faster of the two gradients (drifting Gabor patches) presented sequentially for 300 ms with an inter-stimulus interval of 500ms ^51^. Velocities of the gradients ranged between 6°/s and 10°/s. There were 9 velocity difference levels to modulate the difficulty of the task. 15 trials were presented at each difficulty level. Velocity discrimination thresholds, which corresponds to an accuracy level equivalent to 75% correct for each subject, calculated as an indicator of performance. This task session lasted for about 5 minutes.

### Supplement 5

#### ERP Paradigms and EEG Preprocessing

##### EEG Recording

EEG was recorded using a Compumedics Neuroscan 64 sensor net with nose reference. Impedances were kept below 10 kΩ and data were sampled at 1000 Hz with a bandpass filter of direct current (DC) to 100 Hz.

##### SSVEP Task

Recording conditions, stimulus presentation, and recording equipment were standardized across sites. Participants refrained from smoking one hour prior to testing. Subjects completed a visual paradigm while seated in a sound and electrically shielded booth. Stimuli consisted of 50 trials of a black and white square oscillating at 18.75 Hz in the subject’s central, bilateral, left, or right visual field for 2000 ms (200 total trials pseudorandomally interleaved) with inter-trial intervals of 2.5 seconds. Participants were asked to press a button on a response pad with their left thumb to five red oscillating squares from each condition interleaved into the paradigm in order to encourage task engagement, but those trials were not used in any further analysis.

##### SSVEP Processing

EEG data were pre-processed following previously published methods (Hamm et al., 2014; Ethridge et al., 2015; Parker et al., 2019). Raw EEG data were inspected for bad sensors and artifacts. Bad sensors were interpolated (<5% for any subject) using spherical spline interpolation (BESA 5.3; MEGIS Software, Grafelfing, Germany) and transformed to an average reference. Blink and cardiac artifacts were removed using independent component analysis (EEGLAB 13.6; Delorme and Makeig, 2004). Data were segmented into 3000-ms epochs from 500 ms before stimulus onset to 500 ms post-onset. Data digitally band pass filtered from .5 Hz to 55 Hz (zero-phase filter; roll-off: 6 and 48 dB/octave, respectively). Epochs containing activity greater than 125 µV at any sensor were not included.

##### IAPS Task

The IAPS stimuli included 60 pseudorandomly ordered scenes including 20 unpleasant, 20 pleasant, and 20 neutral stimuli. Scenes consisted of human threat, animal threat, erotica, romantic couples, people, families, and landscapes. During each experimental session, participants viewed each scene three times. Images were presented for 1000 ms and then followed by 3.5 s of a black screen with a small red dot as a fixation point. After the task was completed, participants were asked to self-rate each scene based on arousal and valence using The Self-Assessment Manikin^52^.

##### IAPS Processing

All processing of data was performed through EEGLAB and ERPLAB under Matlab 2019a. Sensors determined to obtain bad recordings were interpolated using a spherical spline method. Data were transformed to an average reference and filtered from 0.5 (12 dB/oct, zero phase) to 50 (48 dB/oct, zero phase) Hz. Data were down sampled to 500Hz. Epochs containing activity greater than 120 μV at any sensor were not included. Eye blink, cardiac, line noise, and muscle artifacts were identified using the Independent Component Analysis (ICA). IClabel was implemented to determine bad components. Components determined to be more than 80 percent artifact, or a bad component were first inspected and then removed if considered to be an artifact.

A baseline correction was performed at -500 before stimulus onset. Out of a possible 60 trails per scene, a minimum of 48 trials were included in each subject’s ERP waveform average.

##### Sensor Selection

Sensor selection to quantify EPNs was performed through visual inspection of topographic activity. Visual inspection took place through mapping of topographic time-lapsed data through EEGLAB To represent the EPN, sensors P7, P8, PO7, PO8, O1, and O2 were averaged over 150–250 ms.

### Supplement 6

#### Primary Outcomes - Supplemental

Removal of imputed PANSS General scores in the HD-tDCS condition resulted in effects surviving our significance threshold (*W*=0.37; p=0.08; 95% CI 0.13, 1.00). Pairwise comparisons showed lower PANSS general scores from baseline to day 5 (RBES=0.48; p_fdr_=0.08; 95% CI - 0.22, 0.85). Similarly, scores significantly increased from day 5 to 1-month in the HD-tDCS condition (RBES=-0.44; p_fdr_=0.08; 95% CI -0.84, 0.27).

Removal of imputed PANSS General scores in the HD-tACS condition resulted in significant effects for the pairwise comparisons, which demonstrated lower PANSS General score between 5 day and 1-month (RBES=0.56; p_uncorrected_=0.07; 95% CI -0.36, 0.93) and baseline and 1-month (RBES=0.56; p_uncorrected_=0.10; 95% CI -0.36, 0.93). However, there was no difference in variance when using the Friedman test across all 3 time points (*W*=0.53; p=0.15; 95% CI 0.44, 1.00).

Removal of imputed SSVEP values resulted in similar effects. There were significant differences across sessions for the SSVEP P1 voltage in the HD-tDCS group for bilateral trials at POz (*W*=0.61; p=0.04; 95% CI 0.848, 1.00). HD-tDCS post hoc analyses still showed a significant reduction in voltage for P1 from baseline to 5 day (RBES=0.32; p_fdr_=0.02; 95% CI -0.40, 0.79) and baseline to 1-month (RBES=0.44; p_fdr_=0.02; 95% CI -0.27, 0.84). The SSVEP N1 voltage remained significantly different across sessions in the HD-tDCS group for bilateral POz (*W*=0.84; p=0.01; 95% CI 0.76, 1.00). HD-tDCS post hoc analyses still showed a significant increase in baseline to 5 day (RBES=-0.76; p_fdr_=0.0007; 95% CI -0.94, -0.24) and from baseline to 1-month (RBES=-0.20; p_fdr_=0.07; 95% CI -0.74, 0.50), as well as a significant decrease from 5 day to 1-month (RBES=0.12; p_fdr_=0.004; 95% CI -0.56, 0.70). HD-tACS effects on P1 and N1 were note examined due to the lack of a significant effect on this electrophysiological measure.

#### Secondary Outcomes - Supplemental

Removal of imputed values resulted in GAF scores no longer being significantly different across sessions in the HD-tACS condition (*W*=0.33; p=0.14; 95% CI 0.08, 1.00). Pairwise comparisons showed GAF increased from day 5 to 1-month (RBES=-0.33; P_uncorrected_=0.07; 95% CI -0.87, 0.57) and baseline to 1-month (RBES=-0.33; P_uncorrected_=0.07; 95% CI -0.87, 0.57).

Removal of imputed IAPS values in the HD-tDCS condition resulted in effects surviving our significance threshold for the unpleasant (*W*=0.81; p=0.04; 95% CI 0.75, 1.00), but not the neutral (*W*=0.44; p=0.17; 95% CI 0.25, 1.00) stimuli. Pairwise comparisons were no longer significantly decreased for unpleasant stimuli from baseline to day 5 (RBES=0.62; p_fdr_ = 0.21; 95% CI -0.12, 0.92), but day 5 to 1-month (RBES=0.62; p_fdr_=0.02; 95% CI -0.12, 0.92) and baseline to 1-month (RBES=0.75; p_fdr_=0.007; 95% CI 0.12, 0.95) remained significant. Pairwise comparisons related to neutral stimuli no longer survived multiple comparison correction, but the decrease noted between baseline and 1-month (RBES=0.62; p_fdr_=0.09; 95% CI -0.12, 0.92) remained significant.

## Supplementary Materials

**eFigure 1:**
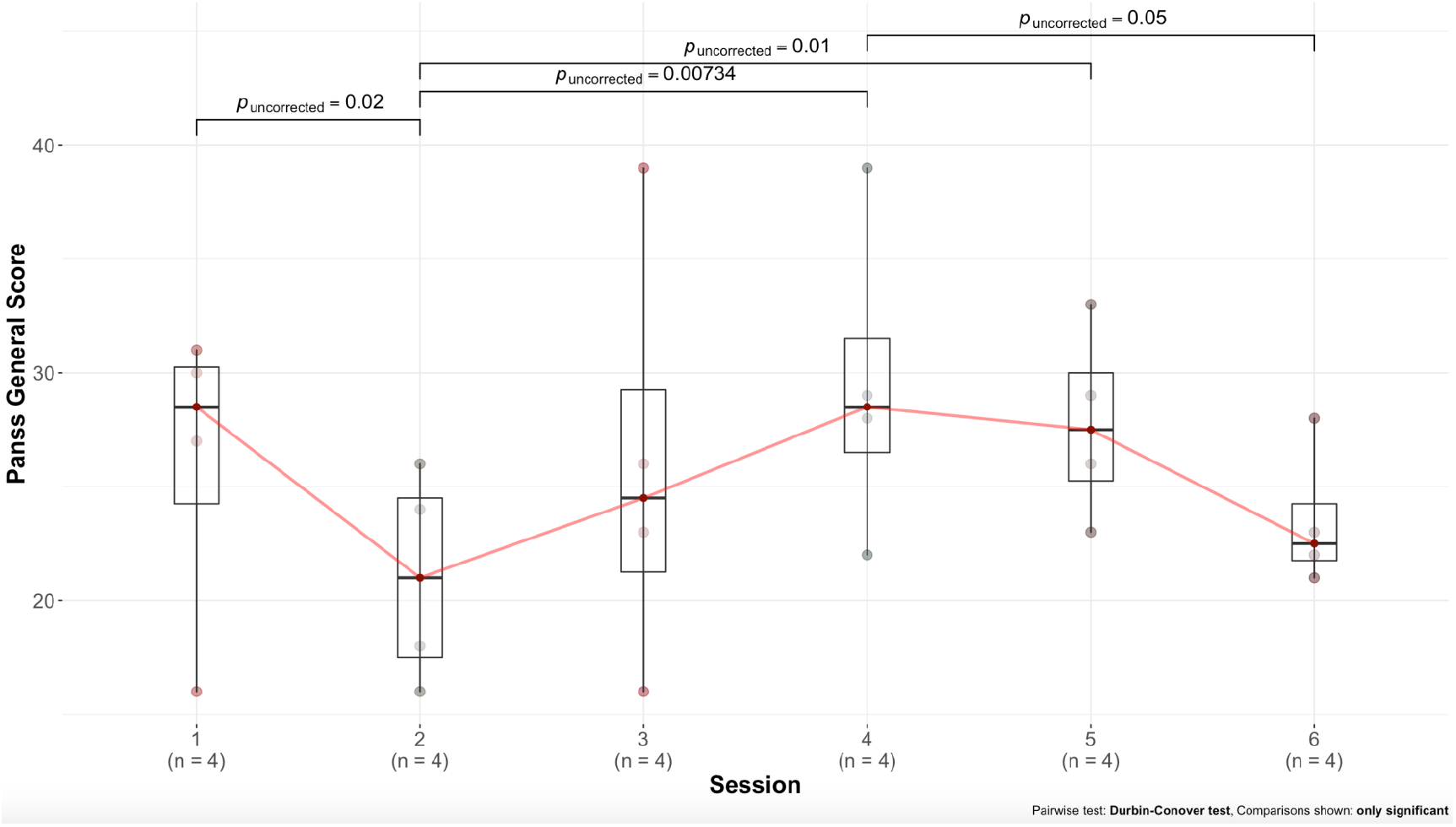
Supplementary Primary Outcome Results: This figure demonstrates the summary of post-hoc pairwise comparisons by session contrasts for both HD-tDCS and HD-tACS in one graph. The purpose for this analysis is to determine whether there was a see-saw effect for PANSS general symptoms by examining differences between HD-tDCS 1-month compared to HD-tACS baseline or between HD-tDCS baseline compared to HD-tACS 1-month.

**eFigure 2:**
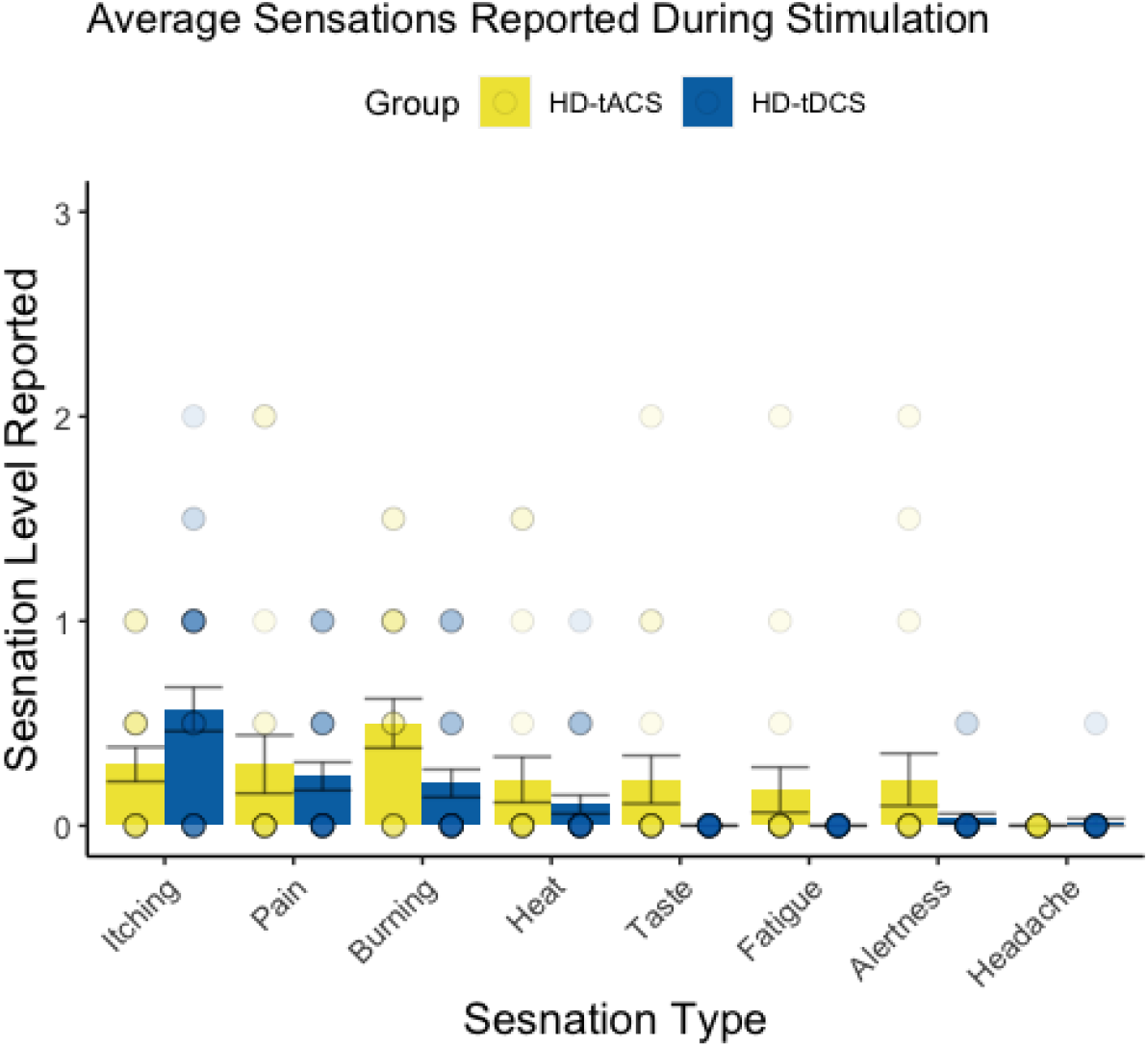
Sensation Questionnaire Results: The graph demonstrates the average sensation (Session 1 and Session 2 of stimulation) reported during HD-tDCS and HD-tACS conditions. Sensations were rated as such: 1, None = I did not feel the sensation addressed; 2, Mild= I mildly felt the sensation addressed; 3, Moderate = I felt the sensation addressed; 4, Strong = I felt the sensation addressed to a considerable degree. **Notes:** HD-tDCS, High-Definition Transcranial Current Stimulation; HD-tACS, HD Transcranial Alternating Current Stimulation.

**eTable 1.** Group by Session Interaction Effects For Outcome Variables

| Table 1. Group by Session Interaction Effects For Outcome Variables |  |  |  |
| --- | --- | --- | --- |
|  | F Value | df1, df2 | P Value |
| **Primary Outcomes** |  |  |  |
| PANSS Total |  |  |  |
| Group | 1.28 | 1, 5.02 | 0.31 |
| Session | 2.85 | 2, 4.17 | 0.17 |
| Group by Session | 4.35 | 2, 4.17 | 0.10 |
| PANSS Positive |  |  |  |
| Group | 0.38 | 1, 5.59 | 0.56 |
| Session | 0.81 | 2, 4.36 | 0.50 |
| Group by Session | 0.12 | 2, 4.36 | 0.89 |
| PANSS Negative |  |  |  |
| Group | 1.05 | 1, 4.36 | 0.36 |
| Session | 0.73 | 2, 3.61 | 0.54 |
| Group by Session | 2.37 | 2, 3.61 | 0.22 |
| PANSS General |  |  |  |
| Group | 1.44 | 1, 5.36 | 0.28 |
| Session | 3.51 | 2, 4.28 | 0.13 |
| Group by Session | **12.42** | **2, 4.28** | **0.02** |
| SSVEP P100 Voltage |  |  |  |
| Group | 0.39 | 1, 4.28 | 0.56 |
| Session | 0.04 | 2, 3.34 | 0.96 |
| Group by Session | 1.37 | 2, 3.34 | 0.37 |
| SSVEP N100 Voltage |  |  |  |
| Group | 0.04 | 1, 5.97 | 0.85 |
| Session | 3.29 | 2, 4.89 | 0.12 |
| Group by Session | 3.21 | 2, 4.89 | 0.13 |
| **Secondary Outcomes** |  |  |  |
| GAF |  |  |  |
| Group | 0.17 | 1, 5.82 | 0.70 |
| Session | 0.14 | 2, 4.31 | 0.87 |
| Group by Session | 1.50 | 2, 4.31 | 0.32 |
| MADRS |  |  |  |
| Group | 0.001 | 1, 5.34 | 0.98 |
| Session | 1.21 | 2, 4.25 | 0.39 |
| Group by Session | 0.77 | 2, 4.25 | 0.52 |
| Model used included Clinical Variable ~ Group * Session, Error = Subject |  |  |  |
| **Notes:** df, Degree of Freedom |  |  |  |

**eTable 2.**
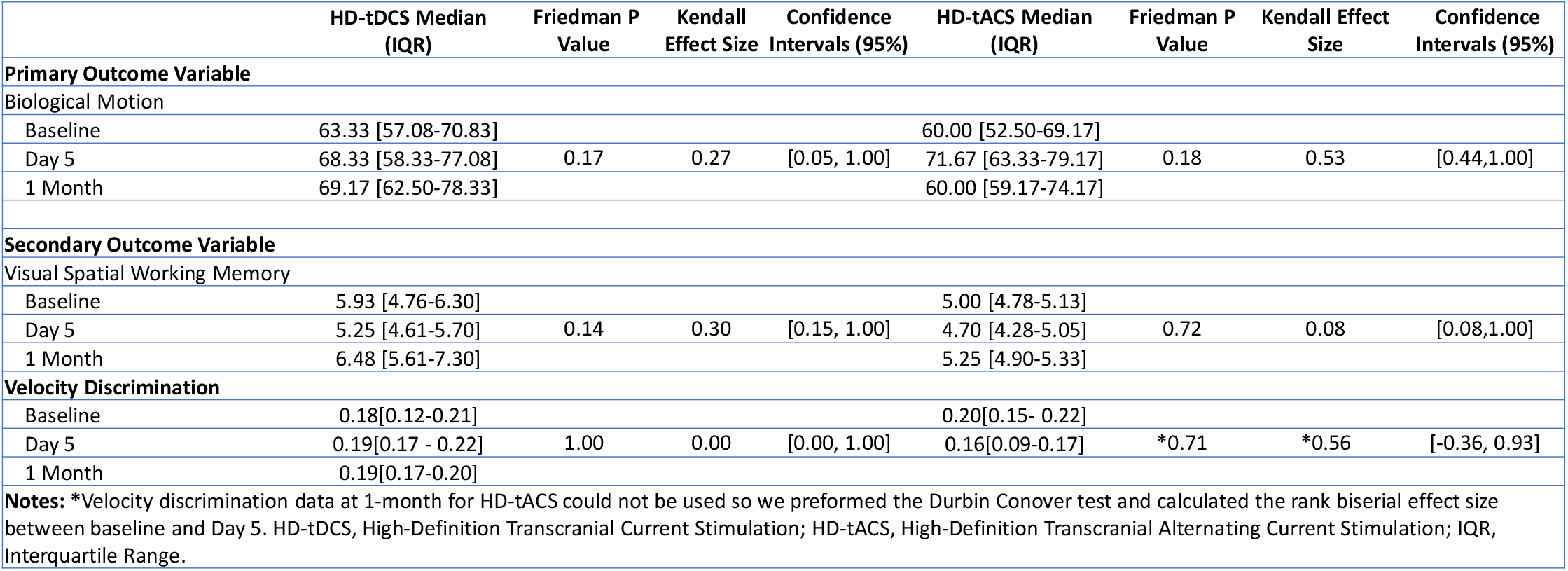
Visual Tasks Outcomes

**eTable 3.** Rank Based Estimation Regression Results Comparing Change from Day 5 to Baseline

|  |  |  |  |  |
| --- | --- | --- | --- | --- |
| | $\beta$ Estimate | SD | T Value | P Value |
| **SSVEP** |  |  |  |  |
| P100~PANSS General |  |  |  |  |
| Intercept | -0.916 | 1.39 | -0.66 | 0.56 |
| PANSS General Change | **0.274** | **0.08** | **3.59** | **0.04** |
| PANSS General Baseline | 0.051 | 0.06 | 0.81 | 0.48 |
| N100~PANSS General |  |  |  |  |
| Intercept | 4.364 | 3.43 | 1.27 | 0.29 |
| PANSS General Change | -0.022 | 0.19 | -0.12 | 0.91 |
| PANSS General Baseline | -0.106 | 0.15 | -0.69 | 0.54 |
| **IAPS EPN** |  |  |  |  |
| Unpleasant EPN~PANSS General |  |  |  |  |
| Intercept | 0.816 | 4.21 | 0.19 | 0.86 |
| PANSS General Change | 0.529 | 0.24 | 2.18 | 0.16 |
| PANSS General Baseline | 0.027 | 0.20 | 0.13 | 0.91 |
| Neutral EPN~PANSS General |  |  |  |  |
| Intercept | 3.543 | 8.19 | 0.43 | 0.71 |
| PANSS General Change | 0.173 | 0.47 | 0.37 | 0.75 |
| PANSS General Baseline | -0.100 | 0.39 | -0.26 | 0.82 |
| Model used included the EEG Variable ~ PANSS General Change + PANSS General Baseline, Score = Skewed Left. **Notes:** PANSS, Positive and Negative Syndrome Scale; SSVEP, Steady State Evoked Potential; IAPS, International Affective Picture System; EPN, Early Posterior Negativity; SD, Standard Error |  |  |  |  |

